# Multi-level multi-state modelling applied to hospital admission in mexican patients with COVID-19

**DOI:** 10.1101/2021.05.24.21257752

**Authors:** Juan Pablo Diaz Martinez, Karen Janik Orozco Becerril, Marco Antonio Gallegos Herrada, Mayra Alejandra Gutierrez Garcia, Osvaldo Espin-Garcia, Ruth Fuentes Garcia

**Affiliations:** Institute of Health, Policy & Management, University of Toronto; Dalla Lana School of Public Health, University of Toronto; Facultad de Ciencias, UNAM

## Abstract

Since the beginning of the SARS-CoV 2 pandemic, healthcare authorities have made clear that it is crucial to track and identify COVID-19 symptoms and seek medical attention in the presence of the first warning signs, as immediate medical attention can improve the patient’s prognosis. Therefore the present work aims to analyze the risks associated with the time between the patient’s first symptoms and hospitalization followed by death. A cross-sectional study was performed among Mexican population diagnosed with COVID-19 and hospitalized from March to January 2021. Four different Bayesian models were developed to asses the risk associated with different patient trajectories: symptoms-hospitalization and hospitalization-death. Comorbidities that could worsen the patient outcome were included as linear predictions; these analyses were further broken down to the different states of the Mexican Republic and the healthcare providers within. Model III was chosen as the best performance through a validation of leaving one out (LOO). Increased risk for hospitalization was observed at the global population level for chronic renal disease, whereas for death such was the case for COPD and the interaction of diabetes:hypertension:obesity. Our results show that there are differences in mortality between the states without accounting for institution and it is related to the prompt time of death or viceversa. Regarding the 6 healthcare providers included in the analysis differences were also found. While state-managed hospitals and private sector showed lower risks, in contrast the IMSS seems to be the one with the highest risk. The proposed modelling can be helpful to improve healthcare assistance at a regional level, additionally it could inform statistical parameter inference in epidemiological models.

## Introduction

The severe acute respiratory syndrome coronavirus 2 (SARS-CoV-2) pandemic was declared a Public Health Emergency of International Concern on January 30, 2020 by the World Health Organization.The Mexican Health Authorities declared the first lockdown on March 26 with 585 cases and 8 deaths reported for COVID-19 [1]; by the end of the first lockdown (June 5th, 2020) total number of cases and deaths were 110,026 and 13,170, respectively. By November 1, Mexico became the fourth country in number of deaths of SARS-CoV-19 (106,765 deaths), with 1,122,362 incident cases [2]; by April 15th 2021 the number of deaths had raised to 214,372 with 2,309,099 incident cases [3].

Over time it has become clear that comorbidity factors such as hypertension, type 2 diabetes mellitus, obesity and smoking increase the seriousness of the disease, leading to a higher rate of hospitalizations with an additional 25% of the cases requiring intensive care unit (ICU) admission and ultimately, intubation and death [4,5].

Mexico ranks second in obesity among OECD countries, with an obesity rate of 72.5% among the adult population, which is associated with the high prevalence of type 2 diabetes mellitus, estimated at 13% of the adult population in 2017, the highest rate among OECD countries; the rate of hypertension is also one of the highest chronic diseases among adult population with 30% [6]. The high prevalence of these comorbidities besides the precarious healthcare system could be among the main reasons of the elevated severity of the number of cases and deaths rates in the country [7,8].

In Mexico, healthcare providers are divided in public and private services. There are different public institutions which provide care to different sets of the population: the state employees (ISSSTE), the army (SEDENA) and naval members (SEMAR), the oil state company (PEMEX) employees and private companies employees (IMSS). There are also public hospitals for population with no health service coverage (SSA adn). In general, the care withing different healthcare providers cannot be considered homogeneous, therefore it should be relevant for the final outcome of a COVID-19 patient.

There have been many efforts using local data to understand how patients with comorbidities are affected by COVID-19; the work by Bello-Chavolla et al. [9] proposed a clinical score to predict COVID-19 lethality, including different factors like type 2 diabetes mellitus and obesity among confirmed and negative COVID-19 cases in Mexico. This work lead to believe that obesity mediates 49.5% of the effect of diabetes on COVID-19 lethality. Also, early-onset diabetes conferred an increased risk of hospitalization while obesity increased the risk ICU for admission and intubation. Moreover, Olivas-Mart {‘i}nez et al. [5] found that main risk factors associated with in-hospital death were male sex, obesity and oxygen saturation *<* 80% on admission using data from a SARS-CoV-2 referral center in Mexico City.

After onset of infection there is a period of time between symptom detection and hospitalization. The time elapsed before patients approach hospitals could be excessively long. Once patients are admitted to hospital, there is also a period of time between the admission and death. Estimation of these lengths of time through a multilevel model could enable a better information system to estimate incidence and transmission rates, particularly at regional level where differences can be apparent. In addition, these times are useful for estimating hospitalisations and deaths in COVID-19 epidemiological models [10,11].

This work considers a multi-state model under a Bayesian framework to estimate times between symptom detection and hospitalization and between hospitalization and death. We used data of confirmed and negative COVID-19 cases and their demographic and health characteristics from the General Directorate of Epidemiology of the Mexican Ministry of Health; the analysis provides of general overview of these times in each state of the country and the different health institutions within. Variables affecting the patient’s final outcome such as the aforementioned comorbidites are included in the model as fixed effects. Additionally, regional heterogeneity is accounted for as random effects through nested models that consider the regional contribution and also the health service provider. Other efforts in recent literature [12] have considered more states (hospitalization-ICU, ICU-death, ICU-discharged), which allows researchers to asses whether improvements in patient outcomes have been sustained, finding evidence that median hospital stays have lengthened. Unfortunately, data available for Mexico lacks the necessary granularity to determine such states. Nevertheless, we believe this model could better inform the estimation of the incidence and transmission rates, which is particularly important while new variants and increased transmission rates are present.

## Methods and materials

### Data Source and Study Population

We conducted an observation study using the official database from the Mexican Ministry of Health, these data provide an overview of hospital admissions, deaths and the period of time between hospitalizations and first COVID-19 symptoms between March and January 2020. The data analyzed included confirmed individuals with a positive test for SARS-CoV-2 [3]; in late 2020, the Mexican Ministry of Health change its confirmed definition with the objective of including postmortem cases. The information recorded on every individual includes: sex, age, nationality, place of residence, migratory status, and different comorbidites. Data registered on the COVID-19 event includes: type of first contact medical unit, management received (either hospitalization or outpatient), and dates of onset of COVID-19 symptoms, admission to hospitalization, and death. Data on the evolution during the stay in the medical units were not released for public use, such as date of recovery. Exclusion criteria were the observations with incomplete data about hospital admission, symptoms or comorbidities. Additionally, patients whose time of initial symptoms was captured as the day they were admitted to hospital were removed, since this time was likely to be unknown. Finally, we only included patients who experienced either hospitalization or death due to the lack of date of recovery in the dataset.

The following variables were included as linear predictors for modeling time from symptoms to hospitalization: presence of chronic obstructive pulmonary disease (COPD), obesity, chronic kidney disease (CKD), asthma and immune-suppression. For time from hospitalization to death, we included the next variables: presence of type 2 diabetes mellitus, COPD, obesity, hypertension, CKD and the interaction between obesity, diabetes and hypertension. Both times also included age and sex as predictors.

About 87% of the population in Mexico is affiliated to some healthcare provider, but during this pandemic the mexican government has established a list of hospitals designated to treat COVID-19 patients without any affiliation distinction. In this study we identified 6 different healthcare providers which were classified according to their sectors IMSS, ISSSTE, SEDENA/SEMAR/PEMEX, SSA, ESTATALES (healthcare provider within each state) these 5 are public care providers while the sixth sector is private hospitals. It is worth mentioning that following the national hospital transformation plan [13], IMSS alone has transformed about 260 medical units to treat COVID-19 patients [14].

### Modelling

We developed four different models for the trajectories of interest, *Symptoms-Hospitalization* and *Hospitalization-Death*.

We used a **QR** reparameterization for the predictor matrix **X**, i.e. **X** = **QR**, where **Q** is an orthogonal matrix and **R** is an upper triangular matrix. This parameterization is recommended when no prior information is available on the location of the predictors’ coefficients [15]. Moreover, we used a noncentered parameterization [16] by shifting the data’s correlation with the parameters to the hyperparameters.

Each model captures different levels of information, as more levels were included it was possible to differentiate the results according to the added information.

#### Model I: One level

Let *M* and *H* correspond to survival times for deaths and hospitalizations, respectively. We assumed that these times are observations from two independent Weibull distributions, such that,

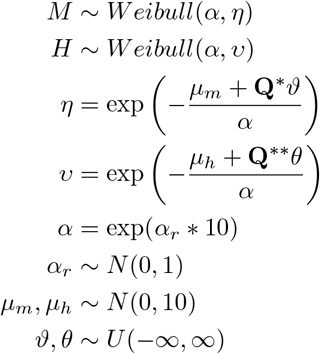

where **Q**^*^ and **Q**^**^ are the orthogonal matrices from the **QR** reparameterization, *θ* and *ϑ* are the coefficient vectors for deaths and hospitalizations, *µ*_*m*_ and *µ*_*h*_ represent the global intercepts for deaths and hospitalizations, *alpha* denotes the shape of the Weibull distribution, and *α*_*r*_ is an extra parameter for the noncentered parameterization This part of the model is described in red in Figure 1.

**Figure 1.**
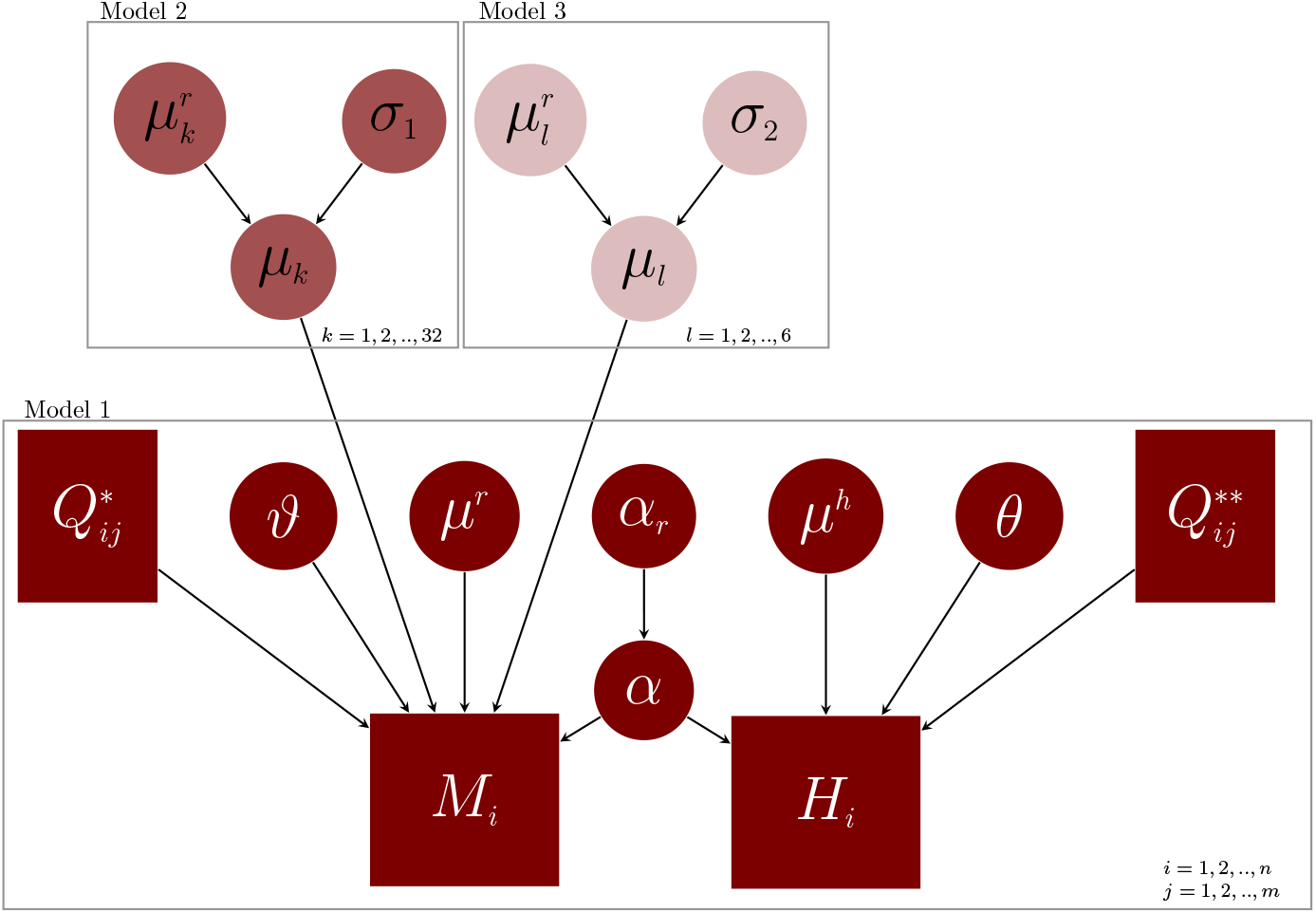
Graphical representation of the model: directed acyclic grph (DAG)

#### Model II: Two levels

The second model adds an additional level to account for each state of Mexico as a random effect to explain deaths, such that,

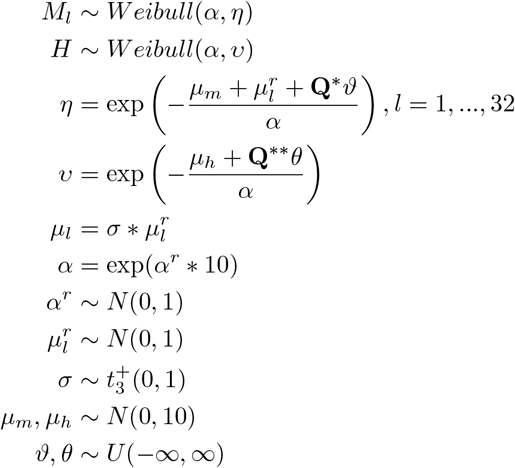

where *µ*_*l*_, *l* = 1, …, 32 represents a local intercept per state (random effect), 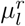 denotes the extra paramateres for the noncentered parameterization, and *σ* is the dispersion parameter of the local intercepts. This part of the model is described in light red in Figure 1.

#### Model III: Three levels

Based on Model II, we considered an extra random effect *µ*_*k*_, *l* = 1, …, 6 which take into account the healthcare provider previously described. Because of this, an extra dispersion parameter *σ* is added. This part of the model is described in light pink in Figure 1.

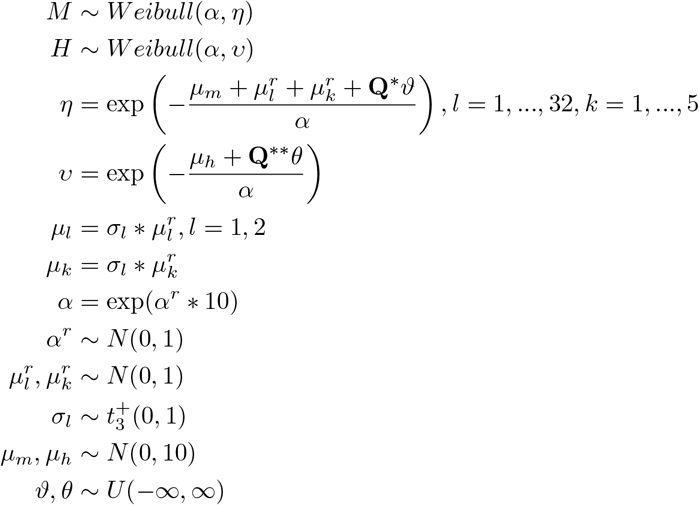

#### Model IV

The final model uses the same structure as Model III but nests healthcare provider within each state.

### Model fit

As mentioned before, we considered a Bayesian approach to fit the model to the observed data corresponding to times between symptoms and hospitalization, *H*, and time between hospitalization and death, *M*. The associated model parameters are given by 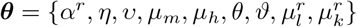. We form the joint posteriordistribution over both the model parameters and auxiliary variables, given the observed data:

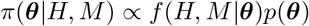

Inference is carried out using Markov chain Monte Carlo (MCMC), obtaining a sample from the joint posterior distribution over the parameters given the observed data. All Markov chains were generated with `CmdStan` [17], using the No-U-Turn sampler [18].

A check on the posterior predictive distributions is essential for validating results. Additionaly, to choose the model that best fits the data we considered the leave-one-out cross-validation (LOO) proposed by Vehtari et al. [19], which estimates pointwise out-of-sample prediction accuracy, using the log-likelihood evaluated at the posterior simulations of the parameter values.

## Results

After applying exclusion criteria a total sample of 1200 registers of adult patients belonging to any healthcare provider, either private or public in the 32 states of Mexico was selected, preserving the population characteristics.

**Table 1.** Expected log-pointwise predictive density (ELPD) for a new data set and effective number of parameters (standard deviation).

| Model | elpd leave one out | p leave one out |
| --- | --- | --- |
| Model I | -75473.3 (187.8) | 26.9 (2.2) |
| Model II | -75352.2 (186.3) | 84.1 (4.7) |
| Model III | -75284.9 (186.6) | 92.5 (4.9) |
| Model IV | 75612.0(194.4) | 239.4(15.0) |

We show results for model III which performed better in terms of the likelihood and showed good convergence of all parameteres. The posterior 0.95 credibility intervals for parameters of interest at different levels of the model are shown in Figures 2 and 3. It is worth pointing out that we are displaying the log hazard ratio, hence positive values for parameters will point to increasing risks for the corresponding transition and level.

**Figure 2.**
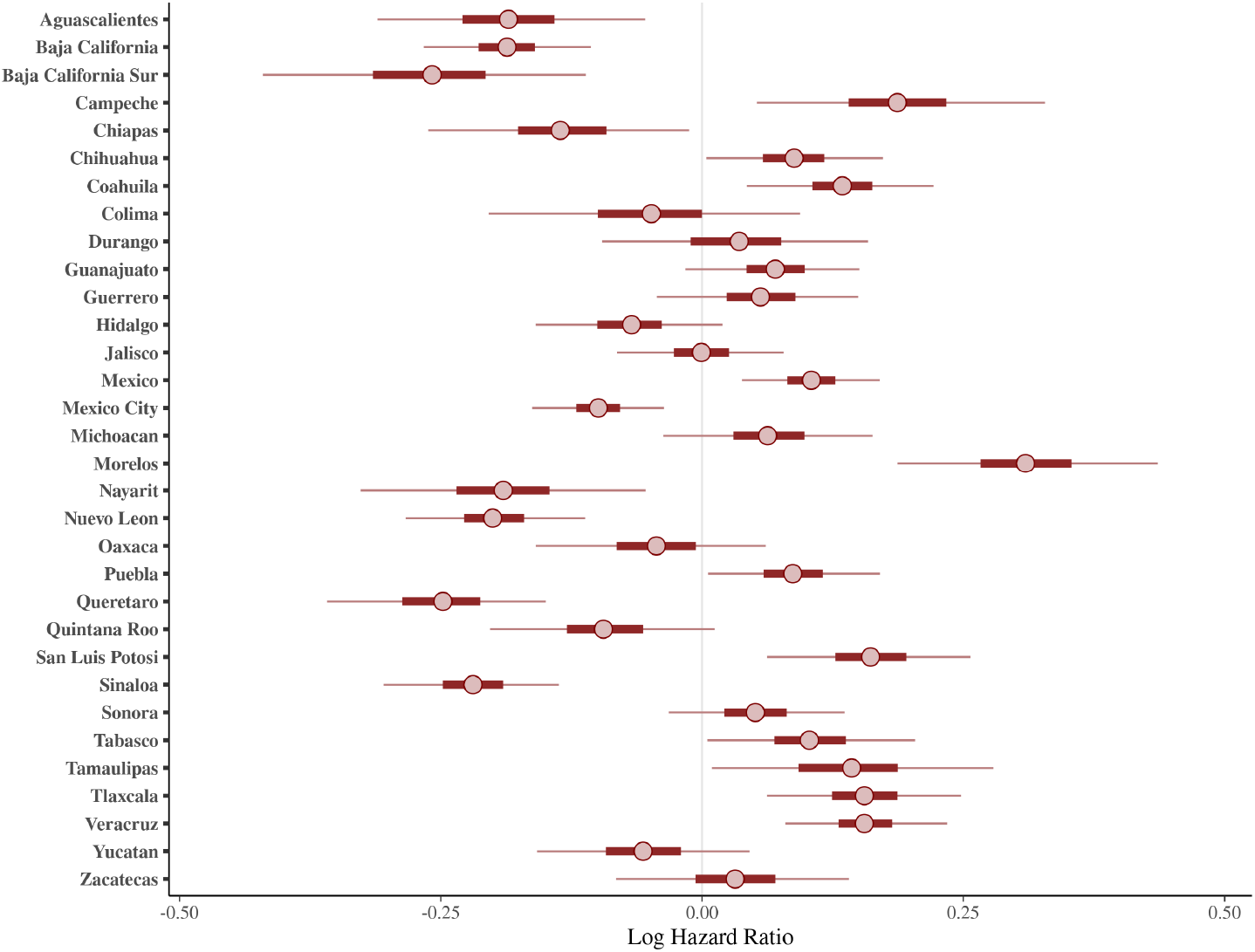
Log hazard rate for state random effect (95% Credible Interval)

**Figure 3.**
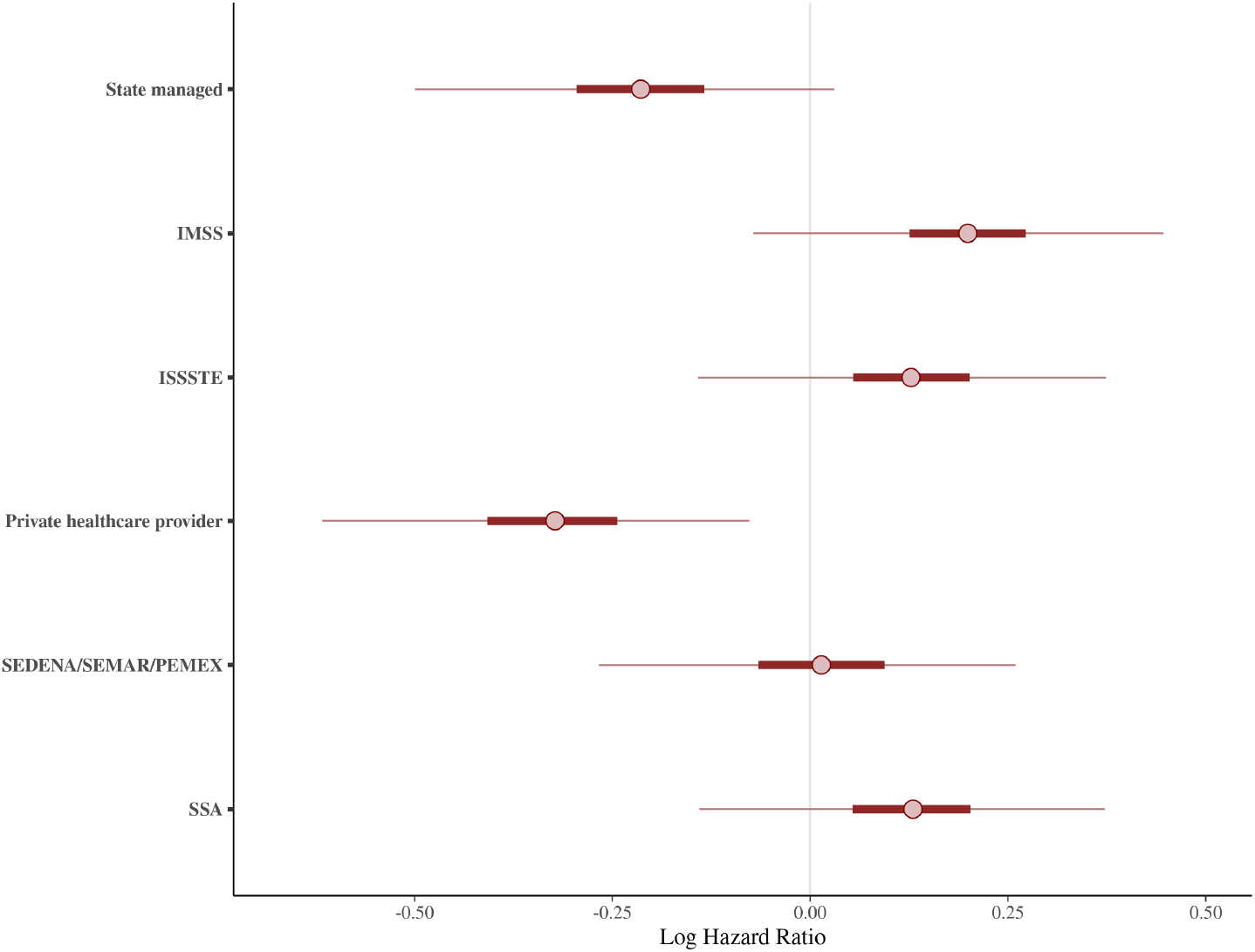
Log hazard rate for healthcare provider random effect (95% Credible Interval)

Increased risk for hospitalization was observed at the global population level for chronic renal disease, whereas for death such was the case for COPD and the interaction of diabetes:hypertension:obesity.

Our results show that there are differences in mortality between the states without accounting for institution and it is related to the prompt time of death or viceversa. Figure 2 shows the states in which the overall rate of mortality is higher such as Campeche, Colima, Guanajuato, Hidalgo, Jalisco, Morelos, Nayarit, Oaxaca, Puebla, Tabasco and Veracruz. The difference might be linked to the late hospitalization of patients.

Figure 4 displays evidence that 5 days after hospitalization there is a peak on mortality rate, which could be related due the late hospitalization of patients with mild symptoms who developed “happy hypoxemia,” that is extremely low blood oxygenation, but without sensation of dyspnea [20]. In Wuhan, within a cohort of patients infected with (SARS-COV-2) who id not present dyspnea 62% showed severe disease and 46% ended up intubated, ventilated or dead [21].

**Figure 4.**
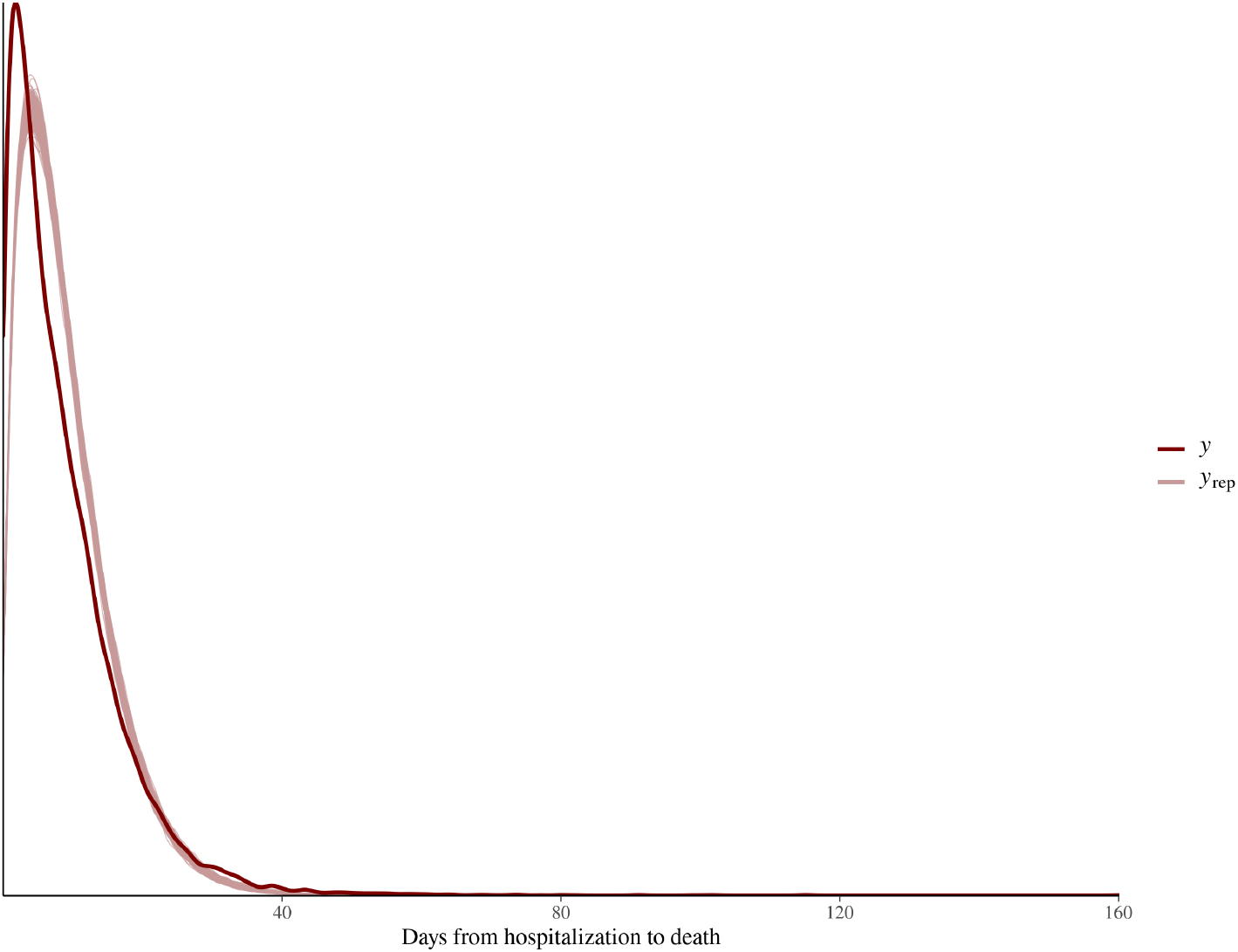
Posterior predictive distribution density plot for deaths

Regarding the 6 healthcare providers included in the analysis differences were also found. While state-managed hospitals and private sector showed lower risks, in contrast the IMSS seems to be the one with the highest risk (Figure 3). Although it is worth mentioning that following the national hospital transformation plan [13], IMSS alone has transformed about 260 medical units to treat Covid-19 patients [14]. Additionaly it has the largest number of affiliations and they are liklely to have higher risk exposure.

## Disscusion

Multiple sources have shown that the presence of comorbidities such as diabetes, hypertension, obesity, chronic obstructive pulmonary disease (COPD), asthma, inmmuno-suppression and chronic kidney disease are associated to a worse outcome for the patients diagnosed with COVID-19. Particularly for those who are hospitalized in the ICU due intubation. To our knowledge this is the first study in Mexico which analyzed the time elapsed between the patient’s first symptoms, hospitalization and death; these analyses were further broken down to the different states of the republic and the healthcare providers within them. Thus it is possible to identify providers and states with an increased risk of hospitalization and death. Mexico is the third place in obesity among the OCDE countries, which could be the main reason of the high number of severe cases of COVID-19.

One problem that aggravates the situation is the precarious state of the public healthcare system which universal coverage is estimated about 87% of the mexican population. It is clear that mexican healthcare has overrun during this pandemic and has appealed to private health providers to cope with the treatment of COVID-19 patients. Among all health care providers the IMSS is the one with the highest risk, however it is the largest healthcare provider across Mexico with hospitals from level 2-level 4 of which “Siglo XXI” is a country-leader in research and innovative treatments and procedures. In March, 2020 38 hospitals were “converted” [13] to exclusively treat COVID-19 among which 18 were IMSS hospitals only; after one year 960 hospitals across the country were converted to treat patients with COVID-19 of these 289 (30.10%) belong to IMSS [14].

The proposed modelling can be helpful to a regional level to improve healthcare assistance, it could additionally be useful to inform statistical estimation of parameters for an epidemiological model.

This study has shown that there are differences in mortality between the different states of the republic; there are states in which the overall rate of mortality is higher due the late hospitalization of patients such as Veracruz, Nuevo Leon, San Luis Potosí, Guanajuato, Chiapas and Mexico City. Breaking down this analysis to state level we found a higher risk of hospitalization, specifically in Veracruz which has been historically unsteady regarding the public healthcare system in sectors like IMSS, ISSSTE, SEDENA/SEMAR/PEMEX. The fact that different final outcomes could be related to patient’s late hospitalization, hence suggesting that the average patient waits until the symptoms are severe to seek professional healthcare, needs to be further investigated.

One of the limitations of the study is the reduced number of states we were able to include in the modls due to the lack of information regarding dates of disccharged of recovered patient’
ss after hospitalization.

## Data Availability

Information is taken from the Mexican Ministry of Health (SISVER).

https://datos.gob.mx/busca/dataset/informacion-referente-a-casos-covid-19-en-mexico

## Author Approval

All authors have seen and approved the manuscript.

